# Atherosclerotic plaque epigenetic age acceleration is characterized by mesenchymal reprogramming and poor prognosis

**DOI:** 10.1101/2023.02.16.23286067

**Authors:** Robin J. G. Hartman, Ernest Diez Benavente, Lotte Slenders, Arjan Boltjes, Barend M. Mol, Gert J. de Borst, Dominique P. V. de Kleijn, Koen H. M. Prange, Menno P. J. de Winther, Johan Kuiper, Mete Civelek, Sander W. van der Laan, Steve Horvath, Charlotte Onland-Moret, Michal Mokry, Gerard Pasterkamp, Hester M. den Ruijter

## Abstract

Epigenetic age estimators (clocks) are known to be predictive of human mortality risk. However, it is not yet known whether the epigenetic age of atherosclerotic plaques can be used for predicting secondary events. Here we estimated an age adjusted measure of epigenetic age, epigenetic age acceleration (EAA), using DNA methylation of human atherosclerotic plaques and of blood. EAA of plaque, but not blood, independently predicted secondary events in a 3-year follow-up (HR=1.3, p= 0.018). Plaque EAA concurred with a high metabolic epigenetic and transcriptional state in plaques. Patients with diabetes and a high body mass index had a higher plaque EAA. EAA was lower in female plaques compared to male plaques by approximately 2 years. Single-cell RNA-seq revealed mesenchymal smooth muscle cells and endothelial cells as main drivers of EAA. Plaque-specific ageing may help identify processes that explain poor health outcomes.

---

Chronological age is a strong predictor for morbidity and mortality. However, people age differently, which matured the concept of epigenetic age. The epigenetic age of a tissue can differ from the chronological age of the same individual. Biological age can be determined in multiple ways, one way is using the Horvath pan tissue clock^1^ as an indicator for epigenetic age. In this method DNA methylation (DNAm) levels at 353 CpG sites is used to predict chronological age. The predicted age estimate is referred to as epigenetic age or DNAm age. An age adjusted measure of DNAm age, known as epigenetic age acceleration, is heritable (up to 40% in adults)^2^ and predicts the prevalence and incidence of many leading diseases^3^. In addition, epigenetic age acceleration (EAA) has been associated with poorer cardiovascular health^4^.

Atherosclerosis is the process in which lipids and immune cells accumulate and form a plaque in the sub-endothelial layer of the blood vessels^5^. This plaque buildup underlies most cardiovascular diseases and is a long-term process that progresses during ageing. We therefore hypothesize that plaque-specific epigenetic ageing measured through DNA methylation reflects progression of disease beyond chronological age.

Histology and imaging studies reveal that overall plaque burden is higher in men compared to women of similar age, and plaque phenotypes in men are associated with more inflammation and an “unstable” phenotype.^6^ With the well-known effects of sex hormones on epigenetic ageing,^7,8^ we postulate that atherosclerotic plaques in women show delayed epigenetic ageing compared to men.

We determined the epigenetic age of 485 atherosclerotic plaques and corresponding blood samples using DNA methylation data. We evaluated the effect of epigenetic age, sex, and other risk factors on follow-up event rates of patients. Lastly, we studied the histological and molecular phenotypes underlying the epigenetic age acceleration of plaques, using histopathological assessment, epigenome-wide DNA methylation signatures and (single-cell) RNA-sequencing. In advanced atherosclerotic disease patients, EAA of the plaque was marginally lower in women compared to men and associated with poorer clinical outcome. Plaque-specific EAA resulted in a large shift in genome-wide epigenetic and transcriptional states which pointed to mesenchymal reprogramming of smooth muscle cells and endothelial cells. Our data highlight the importance of epigenetic aging in driving cardiovascular disease.

## Methods

### Study population

The Athero-Express is an ongoing, prospective biobank study, collecting atherosclerotic plaques from patients undergoing carotid endarterectomy (CEA) in two Dutch tertiary referral hospitals: University Medical Centre Utrecht and St Antonius Hospital, Nieuwegein. Study design, inclusion criteria have been published previously^19^. A baseline table for the 485 patients (148 women and 337 men) in this study is provided in Supplemental Dataset 1. Indication for surgery was based on international guidelines for carotid and iliofemoral atherosclerotic disease^20–23^, and standardized treatment protocols and operative techniques were applied. The medical ethics committees in both participating centres approved the study. All patients provided written informed consent. A composite cardiovascular endpoint for the outcome analysis of patients who underwent CEA was used. This consisted of: (sudden) cardiovascular death, stroke, myocardial infarction, coronary intervention (coronary artery bypass grafting or percutaneous coronary intervention), peripheral (re)intervention or leg amputation. For patients who reached multiple endpoints during follow-up, only the first manifestation of a cardiovascular event was used for analysis of the composite endpoint.

### Plaque histopathology

As described previously^19,24,25^, the atherosclerotic plaque was processed directly after surgery and (immune-)histochemical staining was routinely performed on the culprit lesion (segment with the highest plaque burden) for identification of macrophages (CD 68), calcification (haematoxylin-eosin (HE)), smooth muscle cells (alfa actin), collagen (Picro Sirius red (PSR)), intra-plaque hemorrhage (HE, Elastin von Gieson staining), vessel density (CD34) and fat (PSR, HE).

### Bioinformatics

We used DNA methylation data from 485 atherosclerotic plaques and 93 overlapping blood samples, for which histological procedures, data collection, and normalization procedures were previously published^26^. In brief, DNA was extracted from stored plaque segments and stored blood samples of patients using standardized in-house protocols as described before in Van der Laan et al^27^. DNA purity and concentration were assessed using the Nanodrop 1000 system (Thermo Scientific, Massachusetts, USA). DNA concentrations were equalized at 600 ng, randomized over 96-well plates and bisulfite converted using a cycling protocol, and the EZ-96 DNA methylation kit (Zymo Research, Orange County, USA). Subsequently, DNA methylation was measured on the Infinium HumanMethylation450 Beadchip Array (HM450k, Illumina, San Diego, USA), which was performed at the Erasmus Medical Center Human Genotyping Facility in Rotterdam, the Netherlands. Processing of the sample and array was performed according to the manufacturer’s protocol. We calculated epigenetic age using Horvath’s clock with the R package cgageR (v0.1.0, accessed from: https://github.com/metamaden/cgageR), which uses methylation values at 353 CpGs to generate a continuous value mimicking a person’s epigenetic age.

The dimensionality reduction of 485 patient samples and B-values of the DNA methylation data at 483,731 CpGs was performed using a t-distributed stochastic neighbor embedding with the rtsne package (v0.15). Differential methylation analysis was performed using limma (v3.46.0)^32^. We called a CpG with a Bonferroni-corrected p-value <0.05 significantly differentially methylated. We used ChIPseeker^33^ (v.1.26.2) to annotate the differentially methylated CpGs. Differentially methylated promoter CpGs were enriched using clusterProfiler^30^.

In 420 of these plaques we were able to collect methylation and RNA sequencing data which was processed accpring to previously published methods^28^. Briefly, the raw RNAseq read counts were corrected for UMI sampling:

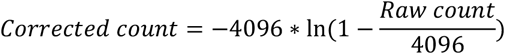

then normalized by sample sequencing depth and log-transformed. Differential gene expression analysis was performed using DESeq2^29^ (v1.30.1). A gene was called differentially expressed if FDR-adjusted p-value < 0.1. Gene enrichment analysis were performed using Clusterprofiler v(3.18.1)^30^, enrichplot (v1.10.2), and enrichr^31^.

### Statistical analysis

All analyses were performed using R (v4.0.4) in R-Studio. Baseline tables were generated using the tableone package (accessed from: https://github.com/kaz-yos/tableone). We calculated the EAA as the residual from a linear model: Epigenetic Age ∼ Chronological Age. This removes the influence of chronological age on epigenetic age, since both are heavily correlated. The same procedure also means that positive EAA values indicate accelerated ageing, whereas negative EAA values indicate decelerated ageing. Linear models were constructed in R adjusting for potential confounders when needed. Cox regressions, plotting, and adjusted cox regressions were performed using rms (v6.2.0), survminer (v0.4.9) and survival (v3.2.10). Code can be found online at www.github.com/

### Single-cell RNA-sequencing

We used previously published single-cell RNA-sequencing data of human atherosclerotic plaques, as well as new data from the same cohort^34^. We were able to distinguish 20 different cell clusters from 46 patients. We used the addModuleScore within seurat^35^ to calculate module scores for genes higher expressed within accelerated plaques, and created violin plots to plot the obtained module scores over the different clusters (Fig. 4C).

## Results

### Chronological age and biological plaque age

To determine the epigenetic age of the atherosclerotic plaque, we used DNA methylation data of plaques from 485 patients (148 women and 337 men) who underwent a carotid endarterectomy. A baseline table with the clinical characteristics of the 485 patients is provided in the Supplemental Dataset 1. Chronological age for the patients ranged from 40 to 91 years in females, and 43 to 88 years in males, with a median age of 69 for both sexes. We used the Horvath pan tissue clock since it applies to all nucleated cells including those found in plaques. Epigenetic age measured using the Horvath pan tissue clock^1^ ranged from 50.4 years of age for females to 81.3 years of age, while it ranged from 51.0 to 88.8 years of age for males, with a median for females of 65.0 years, and 67.6 years of males (Fig. 1A). Chronological age of the patient and epigenetic age of atherosclerotic plaques were strongly correlated (r = 0.695, *p*-value = 3.5e-71, Fig. 1B), indicating that the Horvath clock is relatively accurate in predicting age in diseased atherosclerotic tissue. We calculated epigenetic age acceleration (EAA) by taking the residuals of the linear model in which epigenetic age was predicted by chronological age. It is a mathematical fact that the resulting measure of age acceleration is not correlated with age (correlation zero). A negative EAA means that the chronological age is higher than the epigenetic age of the sample under study, which points to decelerated ageing of the atherosclerotic plaque, whereas a positive EAA indicates accelerated ageing of the plaque. EAA ranged from -15.4 years to 12.6 years in females, and from -10.6 years to 14.4 years in males (Fig. 1C). The median EAA significantly differed between females and males by approximately 2 years (females: -2.2, IQR [-4.3, 2.2] years vs. males: 0.3, IQR [-2.9, 3.8] years; *p*-value: 5.99e-5).

**Figure 1.**
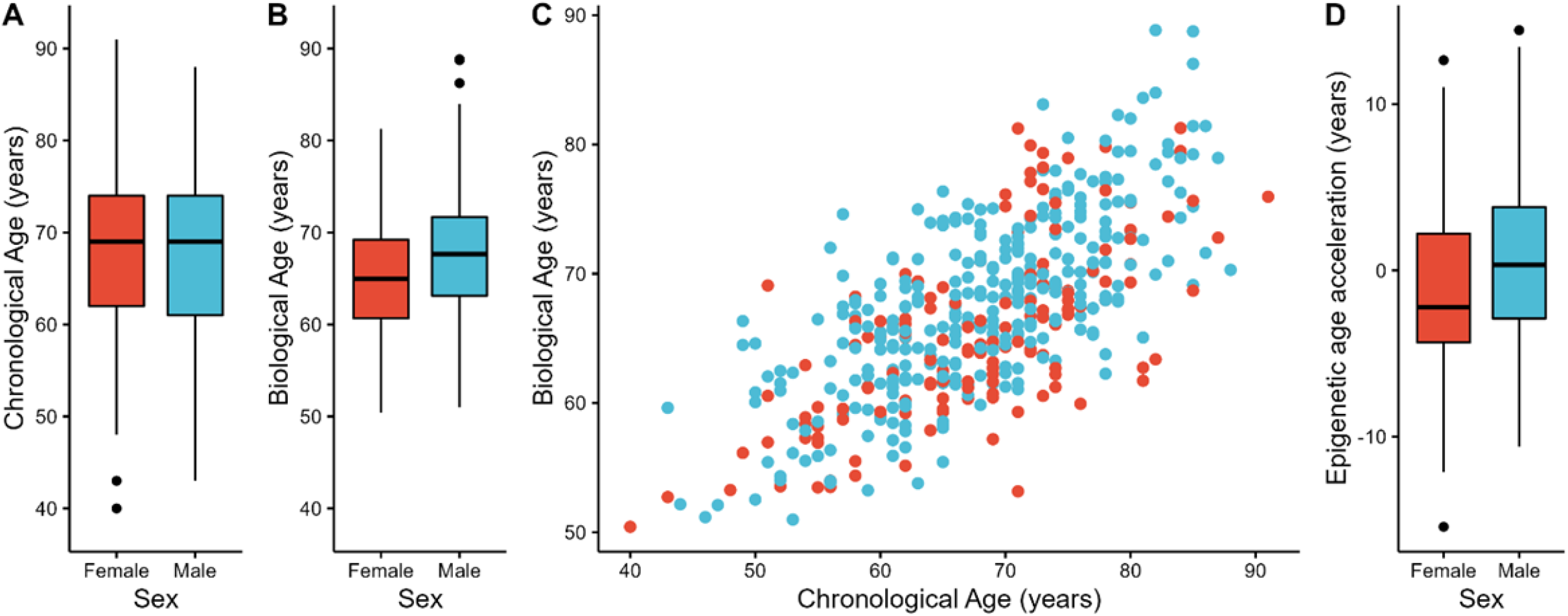
Chronological age versus plaque epigenetic age in 485 carotid endarterectomy patients. A) A boxplot shows the distribution and range of chronological age for the 485 patients in the study, stratified by sex. B) A boxplot shows the distribution and range of epigenetic age determined from DNA of 485 atherosclerotic plaques using the Horvath clock, stratified by sex. C) A scatterplot shows the strong correlation between chronological age (x-axis) and epigenetic age (y-axis), dots are color based on sex. D) A boxplot shows the distribution of range of epigenetic age acceleration (EAA), sex-stratified.

### Epigenetic age acceleration is associated with secondary cardiovascular events

Patients within the Athero-Express biobank are followed up for 3 years for secondary events (see Methods). First, we performed a standard Cox-regression on EAA and secondary events (Fig. 2). To identify any risk factors associated with EAA, we categorized our patient population in two groups, those with accelerated ageing (EAA > 0, n = 224) and those with decelerated ageing (EAA < 0, n = 261) (Table 1). Importantly, since EAA is defined as the residual of the linear model specified above, chronological age is not different between the two groups. Notable differences between the groups are the number of males and females (accelerated: 80% male, decelerated: 60% male, *p*-value < 0.001), their body-mass index (BMI) (accelerated: 27.1 ± 4.2 SD, decelerated: 26.1 ± 3.6 SD, *p*-value = 0.004), and the presence of diabetes (accelerated: 26.8% diabetic, decelerated: 18.4% diabetic, *p*-value = 0.0035). No other risk factors (plasma low-density lipoprotein (LDL), plasma high-density lipoprotein (HDL), hypertension, smoking and high sensitivity C-reactive protein (C-RP)) significantly differed between patients with accelerated and decelerated plaque ageing.

**Figure 2.**
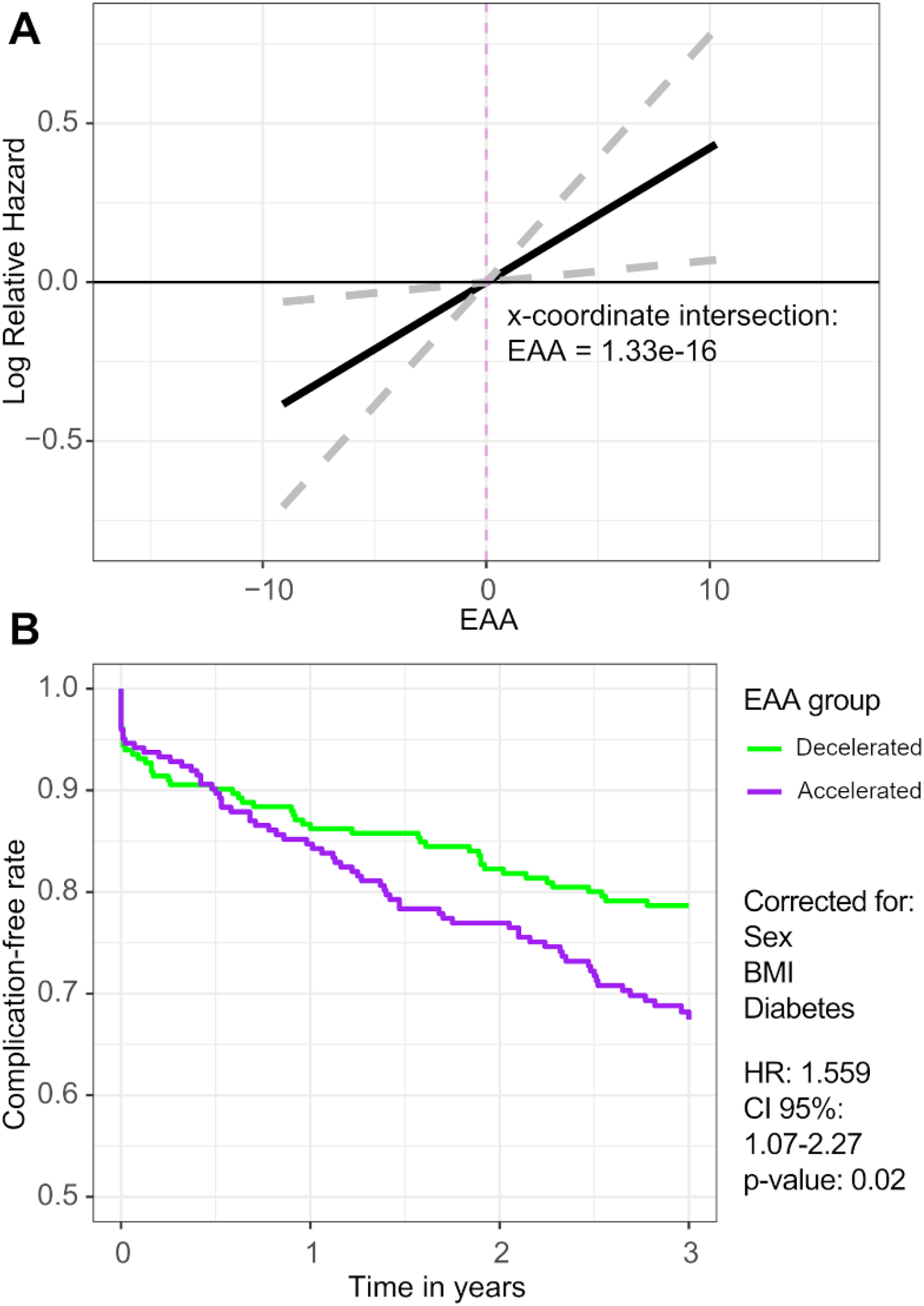
Epigenetic age acceleration of carotid plaques and follow-up in 485 male and female carotid endarterectomy patients. A) A line-graph portrays the relationship between the log of the relative hazard and EAA. The line intersects the y-axis at EAA = 1.33e-16, indicated by the dashed purple line. Dashed grey lines indicate the confidence interval. B) Cox-regression survival graphs, adjusted for sex, BMI, and diabetes, are shown for accelerated (purple) and decelerated (green) plaques.

**Table 1.** Baseline table of patient clinical characteristics of patients with accelerated and decelerated plaques. For continuous variables the value is the average and in parenthesis is the standard deviation (SD), for categorical variables the value represents counts and the parenthesis percentage (%). For non-normally distributed variables (marked with *) the median and the inter-quantile range [IQR] are presented.

| Characteristic | Epigenetic Age<br>Deceleration<br>(N = 261) | Epigenetic Age<br>Acceleration<br>(N = 224) | P value |
| --- | --- | --- | --- |
| Chronological age (years) | 68 (9) | 68 (9) | 0.692 |
| Smoking = yes (%) | 102 (39.7) | 92 (41.8) | 0.705 |
| Sex = male (%) | 158 (60.5) | 179 (79.9) | **<0.001** |
| BMI (kg/m<sup>2</sup>)* | 25.9 [24, 28.3] | 26.5 [24.5, 29] | **0.019** |
| Hypertension = yes (%) | 224 (85.8) | 201 (89.7) | 0.244 |
| Diabetes Mellitus = yes (%) | 48 (18.4) | 60 (26.8) | **0.035** |
| GFR CG (ml/min/1.73 m<sup>2</sup>)* | 72 [59.9, 90] | 73 [53.6, 90] | 0.641 |
| Systolic Blood Pressure (mmHg) | 156 (26.4) | 156 (25.5) | 0.898 |
| Diastolic Blood Pressure (mmHg) | 83 (13) | 82 (13.3) | 0.384 |
| Statin use = yes (%) | 198 (75.9) | 169 (75.4) | 1 |
| Dipyridamole use = yes (%) | 135 (51.7) | 105 (46.9) | 0.33 |
| Aspirin use = yes (%) | 96 (36.8) | 68 (30.4) | 0.163 |
| Contralateral stenosis (%) |  |  | 0.107 |
| 0-50% | 147 (60.0) | 101 (48.6) |  |
| 50-70% | 27 (11.0) | 31 (14.9) |  |
| 70-99% | 29 (11.8) | 33 (15.9) |  |
| 100% | 42 (17.1) | 43 (20.7) |  |
| Symptoms (%) |  |  | 0.587 |
| Asymptomatic | 47 (18.1) | 32 (14.3) |  |
| Ocular | 35 (13.5) | 30 (13.4) |  |
| Transient Ischemic Attack | 115 (44.2) | 98 (43.8) |  |
| Stroke | 63 (24.2) | 64 (28.6) |  |
| Plaque phenotype (%) |  |  | 0.093 |
| Atheromatous | 84 (32.2) | 55 (24.6) |  |
| Fibroatheromatous | 104 (39.8) | 89 (39.7) |  |
| Fibrous | 73 (28.0) | 80 (35.7) |  |
| Intraplaque haemorrhage (%) | 180 (69.2) | 140 (62.5) | 0.143 |
| High-sensitivity C-RP (mg/L) | 6.7 (14.3) | 9.4 (18.1) | 0.231 |
| HDL (mmol/L)* | 1.1 [0.9, 1.4] | 1.1 [0.9, 1.2] | 0.282 |
| LDL (mmol/L)* | 2.6 [2.1, 3.4] | 2.7 [2.2, 3.6] | 0.431 |
| Glucose (mmol/L)* | 5.9 [5.2, 6.9] | 6 [5.3, 7] | 0.595 |

To investigate prognosis in the two EAA-groups we performed a Cox-regression, which showed that being in the group with accelerated plaque ageing was detrimental for secondary outcome (HR = 1.7, *p*-value = 0.0034). This result did not substantially change when we corrected for sex, BMI, and diabetes (adjusted Cox-graphs Fig. 2B, HR = 1.56, *p*-value = 0.02). Adjustment by blood EAA (see Methods) in a subset of patients (n = 93) revealed this to be a plaque-specific effect (HR plaque EAA = 1.3, *p*-value = 0.018; HR blood EAA = 0.1, *p*-value = 0.81), furthermore blood EAA was not predictive for outcome in a separate model (n = 93, HR = 1.6, p-value = 0.44, Supplemental Fig. 1).

Interestingly, we observed a moderate correlation between plaque EAA and blood-based EAA across the subset of patients for whom both measurements were available (correlation coefficient = 0.44, *p*-value < 0.001, n = 93). The moderate correlation illustrates that blood is only a poor surrogate for plaque tissue. There was no significant interaction between EAA and sex (p-value = 0.88). As prognosis in these patients is affected by plaque phenotypes^9^, we studied if EAA was specifically higher in plaques with plaque hemorrhage. However, plaque hemorrhage was not related with EAA, and adjustment for plaque hemorrhage of the Cox-regression showed independent association with secondary outcome for both plaque hemorrhage (HR = 1.6, *p*-value = 0.02) and EAA (HR = 1.7, *p*-value = 0.007).

### Accelerated ageing is independent of plaque characteristics

As plaque accelerated ageing independently predicts cardiovascular secondary outcome, we tested if this coincided with a certain plaque phenotype. The group with accelerated ageing did not present a different profile of semi-quantitative hallmarks of plaque histology: fat, plaque hemorrhage, collagen, calcifications, smooth muscle cell content and macrophage content (see Methods for staining procedures, Supplemental Fig. 2). Plaque phenotype (as shown in Table 1) can be seen as a proxy for plaque cellular composition which can significantly affect epigenetic age calculations^2^. Therefore, we investigated if adjusting the Cox-regression model for secondary cardiovascular outcome by plaque phenotype changed outcome. All analyses remained unchanged and significant, independently of plaque phenotype (HR = 1.6, *p*-value = 0.021). This suggests that EAA predicts secondary cardiovascular outcome independent of histological plaque composition. This prompted us to investigate the effect of EAA on plaque biology at a molecular level by using genome wide epigenetic signatures (beyond the 353 CpGs on which the EAA clock was built on). In addition, we investigated gene expression patterns in accelerated and decelerated plaques.

### Epigenetic age acceleration drives the genome-wide epigenetic state in human plaques

EAA has previously been described as a proxy for the state of the epigenetic maintenance system^1^. An accelerated ageing in our case would indicate a that the epigenetic machinery in the plaque has been under an increased biological workload. In contrast, a decelerated plaque ageing would indicate that the epigenetic machinery has not been under a strong epigenetic maintenance influence as would be expected from plaques which are biologically less active^10^. We therefore studied DNA-methylation patterns in accelerated and decelerated aged plaques (n=485 male and female plaques). For this, we used dimensionality reduction on the DNA methylation data of 483,731 CpGs. We excluded the Horvath-clock CpGs to exclude ageing-related methylation patterns (Fig. 3A). We observed two large clusters representing male and female plaques in our cohort (Fig. 3A). This is likely driven by the differences in DNA methylation of the sex chromosomes: X inactivation leads to hypermethylation of many X chromosomal markers in females ^11,12^. Decelerated and accelerated aged plaque clustered separately within the female, and to some extent, the male plaque groups (Fig. 3A). This suggests that besides sex, EAA is one of the largest drivers of DNA methylation in plaques. Taking male and female plaques together, we found 65,488 differentially methylated CpGs between decelerated and accelerated aged plaques (Supplemental Dataset 2). EAA was associated with hypermethylation of CpGs (45,287 out of 65,488, Fig. 3B). By using gene enrichment analysis on genes marked with differentially methylated CpGs in their promoter (within 1000 bases of the transcription start site), we found enrichment for smooth muscle cell contraction and extracellular matrix organization. We found immune activation and cell adhesion enrichment for hypomethylated promoters of EAA plaques (Fig. 3C). These enrichments were consistent in both females and males when stratifying by sex (Supplemental Fig. 3). We also studied enrichment of atherosclerotic pathways within the Horvath clock CpGs, but no signal was found for immune cell activation and smooth muscle cell pathways. This is in line with previous results that highlighted the processes enriched within the clock CpGs which point to cell death/survival, cellular growth/proliferation, organismal/tissue development, and cancer^2^.

**Figure 3.**
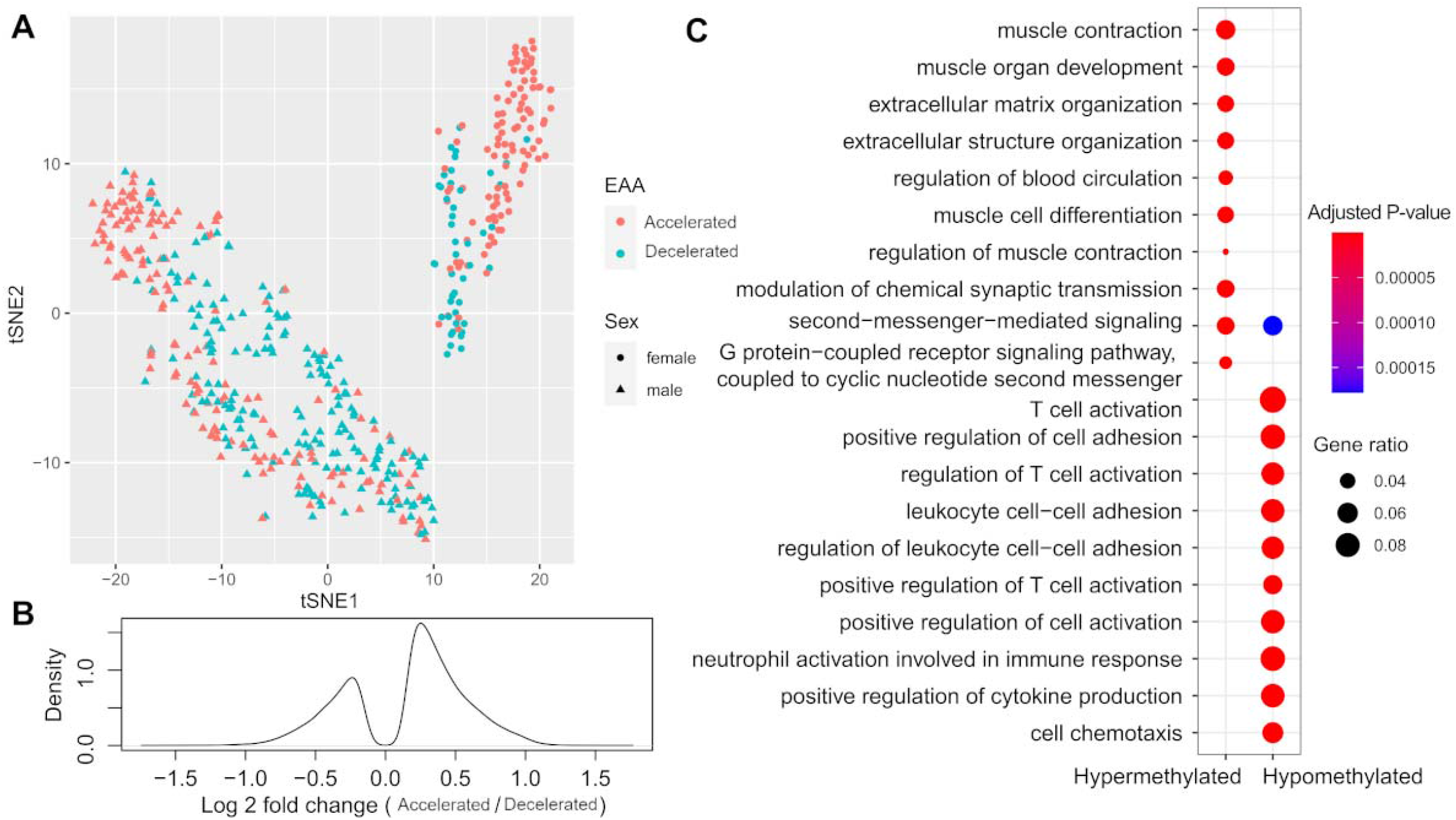
Epigenetic age acceleration and DNA methylome of 485 male and female carotid plaques. A) A tSNE-plot is shown, reducing data of 485 samples based on project 483731 CpGs into 2 dimensions. Shape of the data point indicates sex, color indicates EAA group. We find visual evidence that females cluster into 2 separate groups that can be distinguished by EAA. B) A density plot shows the distribution of log2 fold changes of all significant differentially methylated CpGs between accelerated and decelerated plaques. C) A dot-plot shows the gene ontology Biological Processes enrichment of promoter CpGs significantly hyper- and hypomethylated with accelerated epigenetic ageing corrected for hospital of inclusion and sex. Color indicates significance of the enrichment, whereas size of the dot indicates gene ratio.

### Smooth muscle cells and endothelial cells show transcriptional changes in epigenetically accelerated plaques

To understand which molecular processes are active during plaque EAA, we studied differential gene expression between accelerated and decelerated plaques using whole plaque RNAseq. Of the 19,291 genes tested, 3,031 were differentially expressed (FDR < 0.1) between decelerated and accelerated plaques (Supplemental Dataset 3). The majority of the differentially expressed genes were higher expressed in accelerated plaques (2839 out of 3031) as compared to decelerated plaques (Fig. 4A). We found upregulated genes encoding for ribosomal proteins, as well as mitochondrial genes, in accelerated plaques. This points towards a general increase in translation turn over and mitochondrial respiratory activity within the plaque (Fig. 4B). Gene enrichment analysis using other established databases (i.e. Reactome, KEGG) on the same genes also highlighted *TGFβ* signaling, endothelial to mesenchymal transition and extracellular matrix-related processes (Supplemental Dataset 4) which point towards smooth muscle cells.

**Figure 4.**
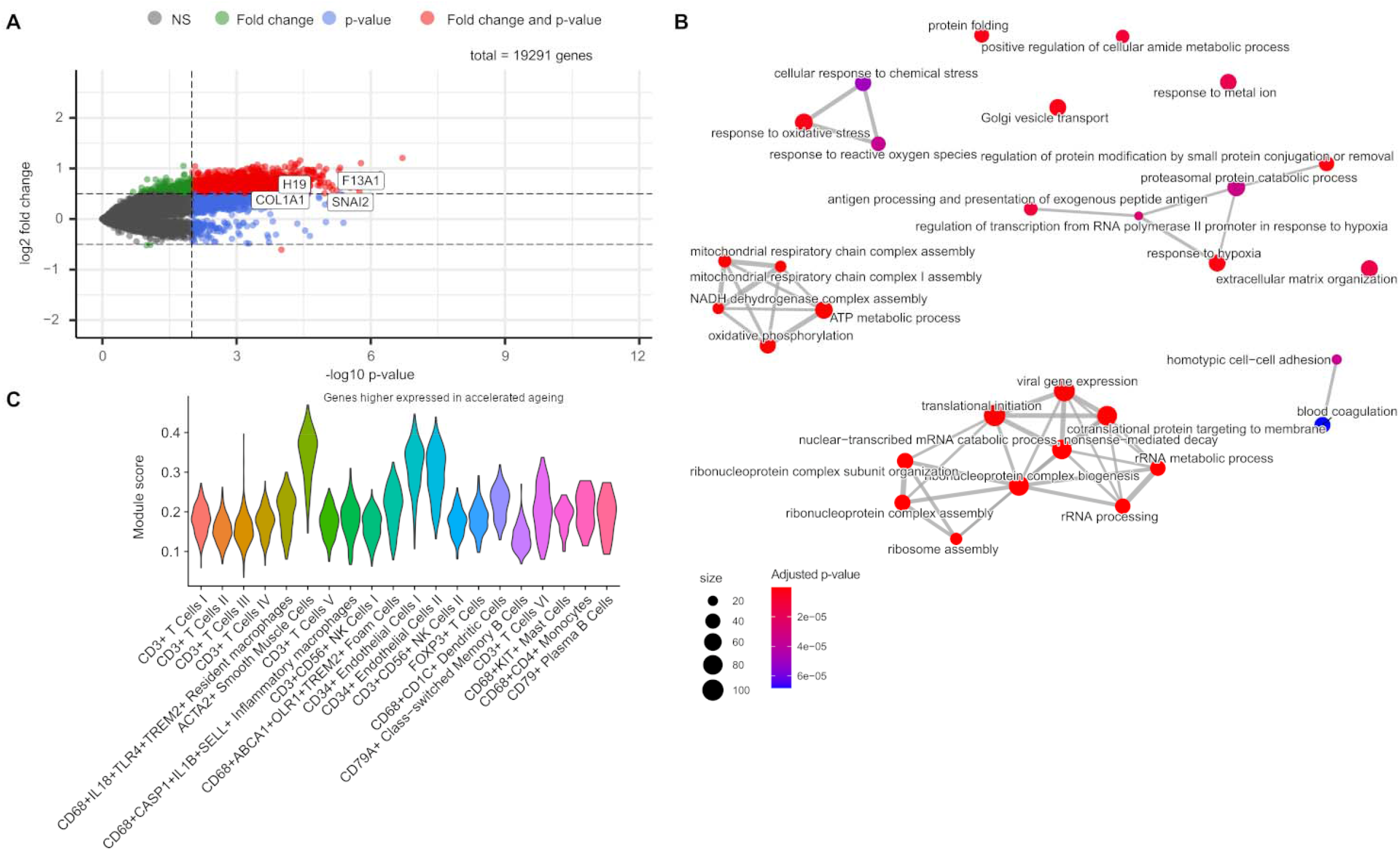
Epigenetic age acceleration and RNA transcriptome of 420 male and female carotid plaques. A) An enhanced volcano-plot is shown for the differential expression analysis between accelerated and decelerated plaques, where the x-axis shows the –log10 of the p-value and the y-axis shows the log 2 fold change (accelerated over decelerated ageing). Notable examples are highlighted in the plot by gene symbol. B) An enrichment map is shown for the gene ontology Biological Processes enrichment performed on genes higher expressed with accelerated ageing. Nodes indicate a gene set, whereas edges indicate overlap between the gene sets. The vicinity of nodes indicates similarity between gene sets. Color highlights significance, whereas size of the nodes indicates the number of genes within that node. C) A violin plot shows the module score of genes higher expressed with accelerated ageing from the bulk RNA-sequencing over the single-cell clusters detected from the single-cell RNA-sequencing of the human atherosclerotic plaque. Colors indicate different cell clusters.

Our transcriptomic and epigenetic analyses of EAA suggest that gene regulation and translation is altered in plaque smooth muscle cells. With the knowledge that smooth muscle cells display remarkable plasticity in atherosclerotic disease, we hypothesized that plaque EAA might alter the transcriptional state of these smooth muscle cells. Therefore, we combined our whole plaque RNA dataset (420 patients, 127 women and 293 men) with single cell RNA data to project the plaque EAA genes over 20 single cell clusters of the plaque including 17 cluster of immune cells (macrophages, T-cells, B-cells, natural killer cells and mast cells) and 3 clusters of mesenchymal cells (*ACTA2*^+^ smooth muscle cells and *CD34*^+^ endothelial cells) within the plaque (4,984 total cells from 46 patients, 20 women and 26 men)^13^. We generated a module score of the genes that were higher expressed in accelerated compared to decelerated plaques and projected this module score onto the individual cells within the 20 identified plaque cell clusters (Fig. 4C). This module score was significantly higher in *ACTA2*^+^ smooth muscle cells (mean module score: 0.15) and the *CD34*^+^ endothelial cell clusters I and II (mean module scores: 0.12 and 0.12, respectively) compared to the rest of the cells clusters within the plaque (mean module score: 0.02; all p-values <0.001) suggesting that mesenchymal cells are most sensitive to accelerated ageing.

## Discussion

Accelerated ageing reflecting that the epigenetic age higher than one’s chronological age, is detrimental for human health (reviewed in^1^). In this study we show that plaque EAA predicts outcome in patients with severe atherosclerotic disease, independent of plaque phenotype. We show that plaques of women are slightly younger compared to men of the same age. We also show that plaque-derived EAA, and not blood-derived EAA, is significantly associated with a poor prognosis in the three years after the initial event. This is independent of other risk factors that are significantly different between patients with accelerated and slowed ageing, such as body mass index and diabetes. Also, plaque phenotypes were not different between those with and without accelerated ageing. Multivariate analysis showed that EAA predicted secondary outcomes in patients independent of plaque hemorrhage. All of this indicates that EAA is a strong biomarker for outcome, independent of the classical plaque paradigm of stable and unstable plaques. Interestingly, plaques from patients with EAA exhibit markedly different DNA methylation patterns, as well as gene expression signatures, compared to patients with decelerated ageing.

We were intrigued to find that the epigenetics ageing and its predictive value was specific to the plaque tissue. Most studies thus far between survival and epigenetic age have been performed on systemic tissue, such as cells from blood. We do not find EAA in blood to be predictive for outcome in our cohort. This highlights that tissue-specificity of the disease is an important characteristic, as described previously^14–16^. This also suggests that EAA in plaque is intimately linked to disease biology within the atherosclerotic plaque. This is supported by the fact that we find ageing acceleration in plaques linked to processes which are important for epigenetic ageing in cells, such as mitochondrial activity and cell plasticity (stem-cell-like potential)^10^.

Whether or not plaque epigenetic age is a proxy for the age of the vasculature that originated the plaque or a proxy for accelerated disease biology in the plaque itself is not known. Cells from the circulation can enter the plaque and become part of the plaque, which may drive some of the effects seen in this study. However, we found that smooth muscle cells and endothelial cells mostly expressed genes that were differentially expressed between accelerated and decelerated plaques. This was not the case for immune cells and suggests that the observed ageing might be predominantly driven by residing cells from the vasculature which were presumably the longest in the plaque environment.

We find a significant sex differences in epigenetic age of around 2 years. Whether or not this is a biological difference or driven by the Horvath clock itself is not known, since previous studies have shown that the obtained epigenetic age is lower in females when using the Horvath clock^7^. It is of interest to note, however, that females develop complications from atherosclerosis almost a decade later in life compared to men^17^. In addition, females outlive males in the human species^18^ and this doesn’t seem to be explained by the epigenetically determined epigenetic age difference. Slower ageing in females as shown by the Horvath clock may be driven by sex differences in gonadal hormones or sex chromosomes and may help explain sex differences in longevity.

We found a marked difference in the genome-wide epigenetic state of accelerated compared to decelerated plaques. When considering genome-wide differentially methylated CpGs between accelerated and decelerated plaques, genes associated with these CpGs were enriched for mesenchymal (hypomethylated) and immune processes (hypermethylated). This was further confirmed by studying differentially expressed genes using RNA-sequencing that pointed towards processes important for inflammation, but also for mesenchymal cells and fibrosis. These results suggest that plaques with accelerated ageing may be more active, with both an inflammatory and mesenchymal component. Taken together, these results point to a combination of these processes being behind the pathophysiological changes occurring during accelerated ageing.

We have shown that positive epigenetic age acceleration in atherosclerotic plaques is associated with a poor prognosis for the patient. Our study has several limitations including the following. First, even though we have acquired RNA and DNA from the same plaques, they originate from different segments of the plaque and small differences might be present. However, the observed results were consistent and in line with known disease mechanisms, therefore, we believe our results are not biased by this. Second, we realize that EAA from plaque is not as easily obtainable as EAA from blood, which may limit the value for clinical practice. Nevertheless, it highlights mechanisms leading to ageing. This might help understand its relevance in disease progression and potentially target it. Lastly, we do not have a point in time which could be used as a reference for when the plaques studied were formed, however we can make inferences from the population and subgroups within the study, which provide relevant information about the processes behind ageing acceleration. Furthermore, by adjusting for chronological age individually and studying the group differences we can get as close as possible to the true epigenetic ageing effect independent of age. We note that the association of blood-derived EAA together with plaque EAA and clinical outcome is based on a smaller sample size. However, it has been reported before that epigenetic age calculation from disease-relevant tissue has better prediction power than that of readily-available tissue^14–16^ and our findings are in line with these reports.

Our results demonstrate that atherosclerotic plaque epigenetic age acceleration independently predicts poor clinal outcome. In addition, female plaques were found marginally biologically younger than male plaques. Plaques from male and female patients with epigenetic age acceleration show a more active and tumultuous epigenetic and transcriptional state, which may be driven by underlying processes in the mesenchymal smooth muscle cells and endothelial cells. These processes may be potential targets for drug discovery and treatment of cardiovascular disease.

## Data Availability

All data and scripts for replication of this study could be provided upon reasonable request.

## List of abbreviations

EAA: Epigenetic age acceleration
EC: Endothelial cell
SMC: Smooth muscle cell

## Acknowledgements

We acknowledge the service of Single Cell Discoveries for single-cell RNA-sequencing of human plaque material.

## Funding

This study was funded by the European Union project European Research Council consolidator grant 866478 (UCARE), and ERA-CVD 2017T099 ENDLESS, Leducq PlaqOmics and AtheroGEN.

## Notes

### Competing Interest Statement

The authors have declared no competing interest.

### Author Declarations

The Athero-Express is an ongoing, prospective biobank study, collecting atherosclerotic plaques from patients undergoing carotid endarterectomy (CEA) in two Dutch tertiary referral hospitals: University Medical Centre Utrecht and St Antonius Hospital, Nieuwegein. Ethical approval was granted by the METC at the University Medical Center Utrecht. Indication for surgery was based on international guidelines for carotid and iliofemoral atherosclerotic disease20-23, and standardized treatment protocols and operative techniques were applied. The medical ethics committees in both participating centres approved the study. All patients provided written informed consent.

## References

1. Horvath S, Raj K. DNA methylation-based biomarkers and the epigenetic clock theory of ageing. Nat Rev Genet [Internet]. 2018;19:371–384. Available from: https://doi.org/10.1038/s41576-018-0004-3

2. Horvath S. DNA methylation age of human tissues and cell types. Genome Biol [Internet]. 2013;14:3156. Available from: https://doi.org/10.1186/gb-2013-14-10-r115

3. Hillary RF, Stevenson AJ, McCartney DL, Campbell A, Walker RM, Howard DM, Ritchie CW, Horvath S, Hayward C, McIntosh AM, Porteous DJ, Deary IJ, Evans KL, Marioni RE. Epigenetic measures of ageing predict the prevalence and incidence of leading causes of death and disease burden. Clin Epigenetics [Internet]. 2020;12:115. Available from: https://doi.org/10.1186/s13148-020-00905-6

4. Pottinger TD, Khan SS, Zheng Y, Zhang W, Tindle HA, Allison M, Wells G, Shadyab AH, Nassir R, Martin LW, Manson JE, Lloyd-Jones DM, Greenland P, Baccarelli AA, Whitsel EA, Hou L. Association of cardiovascular health and epigenetic age acceleration. Clin Epigenetics [Internet]. 2021;13:42. Available from: https://doi.org/10.1186/s13148-021-01028-2

5. Libby P, Buring JE, Badimon L, Hansson GK, Deanfield J, Bittencourt MS, Tokgözoğlu L, Lewis EF. Atherosclerosis. Nat Rev Dis Prim. 2019;5:56.

6. Man JJ, Beckman JA, Jaffe IZ. Sex as a Biological Variable in Atherosclerosis. Circ Res [Internet]. 2020;126:1297–1319. Available from: https://doi.org/10.1161/CIRCRESAHA.120.315930

7. Horvath S, Gurven M, Levine ME, Trumble BC, Kaplan H, Allayee H, Ritz BR, Chen B, Lu AT, Rickabaugh TM, Jamieson BD, Sun D, Li S, Chen W, Quintana-Murci L, Fagny M, Kobor MS, Tsao PS, Reiner AP, Edlefsen KL, Absher D, Assimes TL. An epigenetic clock analysis of race/ethnicity, sex, and coronary heart disease. Genome Biol. 2016;17:0–22.

8. Sugrue VJ, Zoller JA, Narayan P, Lu AT, Ortega-Recalde OJ, Grant MJ, Bawden CS, Rudiger SR, Haghani A, Bond DM, Hore RR, Garratt M, Sears KE, Wang N, Yang XW, Snell RG, Hore TA, Horvath S. Castration delays epigenetic aging and feminizes DNA methylation at androgen-regulated loci. Elife. 2021;10.

9. Hellings WE, Peeters W, Moll FL, Piers SRD, van Setten J, Van der Spek PJ, de Vries J-PPM, Seldenrijk KA, De Bruin PC, Vink A, Velema E, de Kleijn DP V, Pasterkamp G. Composition of carotid atherosclerotic plaque is associated with cardiovascular outcome: a prognostic study. Circulation. 2010;121:1941–1950.

10. Kabacik S, Lowe D, Fransen L, Leonard M, Ang S-L, Whiteman C, Corsi S, Cohen H, Felton S, Bali R, Horvath S, Raj K. The relationship between epigenetic age and the hallmarks of aging in human cells. Nat Aging [Internet]. 2022;2:484–493. Available from: https://doi.org/10.1038/s43587-022-00220-0

11. McCarthy MM, Auger AP, Bale TL, De Vries GJ, Dunn GA, Forger NG, Murray EK, Nugent BM, Schwarz JM, Wilson ME. The Epigenetics of Sex Differences in the Brain. J Neurosci [Internet]. 2009;29:12815–12823. Available from: https://www.jneurosci.org/content/29/41/12815

12. Liu J, Morgan M, Hutchison K, Calhoun VD. A Study of the Influence of Sex on Genome Wide Methylation. PLoS One [Internet]. 2010;5:e10028. Available from: https://doi.org/10.1371/journal.pone.0010028

13. Slenders L, Landsmeer LPL, Cui K, Depuydt MAC, Verwer M, Mekke J, Timmerman N, van den Dungen NAM, Kuiper J, de Winther MPJ, Prange KHM, Ma WF, Miller CL, Aherrahrou R, Civelek M, de Borst GJ, de Kleijn DP V, Asselbergs FW, den Ruijter HM, Boltjes A, Pasterkamp G, van der Laan SW, Mokry M. Intersecting single-cell transcriptomics and genome-wide association studies identifies crucial cell populations and candidate genes for atherosclerosis. Eur Hear J open. 2022;2:oeab043.

14. Levine ME, Lu AT, Bennett DA, Horvath S. Epigenetic age of the pre-frontal cortex is associated with neuritic plaques, amyloid load, and Alzheimer’s disease related cognitive functioning. Aging (Albany NY). 2015;7:1198–1211.

15. Horvath S, Oshima J, Martin GM, Lu AT, Quach A, Cohen H, Felton S, Matsuyama M, Lowe D, Kabacik S, Wilson JG, Reiner AP, Maierhofer A, Flunkert J, Aviv A, Hou L, Baccarelli AA, Li Y, Stewart JD, Whitsel EA, Ferrucci L, Matsuyama S, Raj K. Epigenetic clock for skin and blood cells applied to Hutchinson Gilford Progeria Syndrome and ex vivo studies. Aging (Albany NY). 2018;10:1758–1775.

16. Sillanpää E, Heikkinen A, Kankaanpää A, Paavilainen A, Kujala UM, Tammelin TH, Kovanen V, Sipilä S, Pietiläinen KH, Kaprio J, Ollikainen M, Laakkonen EK. Blood and skeletal muscle ageing determined by epigenetic clocks and their associations with physical activity and functioning. Clin Epigenetics. 2021;13:110.

17. Townsend N, Wilson L, Bhatnagar P, Wickramasinghe K, Rayner M, Nichols M. Cardiovascular disease in Europe: Epidemiological update 2016. Eur Heart J. 2016;37:3232–3245.

18. Jylha J. Sex differences in biological aging with a focus on human studies. 2021;1–27.

19. Verhoeven BAN, Velema E, Schoneveld AH, Paul J, De Vries PM, De Bruin P, Seldenrijk CA, De Kleijn DP V, Busser E, Van Der Graaf Y, Moll F, Pasterkamp G. Athero-Express: Differential Atherosclerotic Plaque Expression of mRNA and Protein in Relation to Cardiovascular Events and Patient Characteristics. Rationale and Design. 2004.

20. Norgren L, Hiatt WR, Dormandy JA, Nehler MR, Harris KA, Fowkes FGR, Bell K, Caporusso J, Durand-Zaleski I, Komori K, Lammer J, Liapis C, Novo S, Razavi M, Robbs J, Schaper N, Shigematsu H, Sapoval M, White C, White J, Clement D, Creager M, Jaff M, Mohler E 3rd, Rutherford RB, Sheehan P, Sillesen H, Rosenfield K. Inter-Society Consensus for the Management of Peripheral Arterial Disease (TASC II). Eur J Vasc Endovasc Surg Off J Eur Soc Vasc Surg. 2007;33 Suppl 1:S1–75.

21. Randomised trial of endarterectomy for recently symptomatic carotid stenosis: final results of the MRC European Carotid Surgery Trial (ECST). Lancet (London, England). 1998;351:1379– 1387.

22. Barnett HJ, Taylor DW, Eliasziw M, Fox AJ, Ferguson GG, Haynes RB, Rankin RN, Clagett GP, Hachinski VC, Sackett DL, Thorpe KE, Meldrum HE, Spence JD. Benefit of carotid endarterectomy in patients with symptomatic moderate or severe stenosis. North American Symptomatic Carotid Endarterectomy Trial Collaborators. N Engl J Med. 1998;339:1415–1425.

23. Halliday A, Mansfield A, Marro J, Peto C, Peto R, Potter J, Thomas D. Prevention of disabling and fatal strokes by successful carotid endarterectomy in patients without recent neurological symptoms: randomised controlled trial. Lancet (London, England). 2004;363:1491–1502.

24. de Bakker M, Timmerman N, van Koeverden ID, de Kleijn Dpv, de Borst GJ, Pasterkamp G, Boersma E, den Ruijter HM. The age- and sex-specific composition of atherosclerotic plaques in vascular surgery patients. Atherosclerosis. 2020;310:1–10.

25. Van Koeverden ID, De Bakker M, Haitjema S, Van Der Laan SW, De Vries JPPM, Hoefer IE, De Borst GJ, Pasterkamp G, Den Ruijter HM. Testosterone to oestradiol ratio reflects systemic and plaque inflammation and predicts future cardiovascular events in men with severe atherosclerosis. Cardiovasc Res. 2019;115:453–462.

26. Siemelink MA, van der Laan SW, Haitjema S, van Koeverden ID, Schaap J, Wesseling M, de Jager SCA, Mokry M, van Iterson M, Dekkers KF, Luijk R, Foroughi Asl H, Michoel T, Björkegren JLM, Aavik E, Ylä-Herttuala S, de Borst GJ, Asselbergs FW, El Azzouzi H, den Ruijter HM, Heijmans BT, Pasterkamp G. Smoking is Associated to DNA Methylation in Atherosclerotic Carotid Lesions. Circ Genomic Precis Med. 2018;11:e002030.

27. Van Der Laan SW, Foroughi Asl H, van den Borne P, van Setten J, van der Perk MEM, van de Weg SM, Schoneveld AH, de Kleijn DPV, Michoel T, Björkegren Jlm, den Ruijter HM, Asselbergs FW, de Bakker PIW, Pasterkamp G. Variants in ALOX5, ALOX5AP and LTA4H are not associated with atherosclerotic plaque phenotypes: The Athero-Express Genomics Study. Atherosclerosis. 2015;239:528–538.

28. Mokry M, Boltjes A, Cui K, Slenders L, Mekke JM, Depuydt MAC, Timmerman N, Waissi F, Verwer MC, Turner AW, Khan MD, Hodonsky CJ, Benavente ED, Hartman RJG, van den Dungen Nam, Lansu N, Nagyova E, Prange KHM, Pavlos E, Andreakos E, Schunkert H, Owens GK, Monaco C, Finn A V, Virmani R, Leeper NJ, de Winther MPJ, Kuiper J, de Borst GJ, Stroes ESG, Civelek M, de Kleijn DP V, den Ruijter HM, Asselbergs FW, van der Laan SW, Miller CL, Pasterkamp G. Transcriptomic-based clustering of advanced atherosclerotic plaques identifies subgroups of plaques with differential underlying biology that associate with clinical presentation. medRxiv [Internet]. 2021;2021.11.25.21266855. Available from: http://medrxiv.org/content/early/2021/11/26/2021.11.25.21266855.abstract

29. Love MI, Huber W, Anders S. Moderated estimation of fold change and dispersion for RNA-seq data with DESeq2. Genome Biol. 2014;15.

30. Yu G, Wang L-G, Han Y, He Q-Y. clusterProfiler: an R Package for Comparing Biological Themes Among Gene Clusters. Omi A J Integr Biol. 2012;16:284–287.

31. Chen EY, Tan CM, Kou Y, Duan Q, Wang Z, Meirelles GV, Clark NR, Ma’ayan A. Enrichr: interactive and collaborative HTML5 gene list enrichment analysis tool. BMC Bioinformatics. 2013;14:128.

32. Ritchie ME, Phipson B, Wu D, Hu Y, Law CW, Shi W, Smyth GK. Limma powers differential expression analyses for RNA-sequencing and microarray studies. Nucleic Acids Res. 2015;43:e47.

33. Yu G, Wang L, He Q. ChIPseeker: an R/Bioconductor package for ChIP peak annotation, comparison and visualization. Bioinformatics. 2015;31:2382–3.

34. Depuydt MA, Prange KH, Slenders L, Örd T, Elbersen D, Boltjes A, de Jager SC, Asselbergs FW, de Borst GJ, Aavik E, Lönnberg T, Lutgens E, Glass CK, den Ruijter HM, Kaikkonen MU, Bot I, Slütter B, van der Laan SW, Yla-Herttuala S, Mokry M, Kuiper J, de Winther MP, Pasterkamp G. Microanatomy of the Human Atherosclerotic Plaque by Single-Cell Transcriptomics. Circ Res. 2020;CIRCRESAHA.120.316770.

35. Butler A, Hoffman P, Smibert P, Papalexi E, Satija R. Integrating single-cell transcriptomic data across different conditions, technologies, and species. Nat Biotechnol. 2018;36:411–420.

